# Modeling the Effect of Population-Wide Vaccination on the Evolution of COVID-19 Epidemic in Canada

**DOI:** 10.1101/2021.02.05.21250572

**Authors:** Soulaimane Berkane, Intissar Harizi, Abdelhamid Tayebi

## Abstract

Population-wide vaccination is critical for containing the COVID-19 pandemic when combined with effective testing and prevention measures. Since the beginning of the COVID-19 outbreak, several companies worked tirelessly for the development of an efficient vaccine that would put an end to this pandemic. Today, a number of COVID-19 vaccines have been approved for use by a number of national regulatory organizations. Vaccination campaigns have already started in several countries with different daily-vaccination rates depending on the country’s vaccination capacity. Therefore, we find it timely and extremely important to conduct a study on the effect of population-wide vaccination campaigns on the evolution of the COVID-19 epidemic. To this end, we propose a new deterministic mathematical model to forecast the COVID-19 epidemic evolution under the effect of vaccination and vaccine efficacy. This model, referred to as SIRV, consists of a compartmental SIR (susceptible, infectious and removed) model augmented with an additional state *V* representing the effectively vaccinated population as well as two inputs representing the daily-vaccination rate and the vaccine efficacy. Using our SIRV model, we predict the evolution of the COVID-19 epidemic in Canada and its most affected provinces (Ontario, Quebec, British Columbia, Alberta, Saskatchewan, and Manitoba), for different daily vaccination rates and vaccine efficacy. Projections suggest that, without vaccination, 219, 000 lives could be lost across Canada by the end of 2021 due to COVID-19. The ongoing vaccination campaign across Canada seems to unfold relatively slowly at an average daily rate close to 1*/*2 vaccine per 1, 000 population. At this pace, we could be saving more than 77, 496 lives by the end of the year. Doubling the current vaccination efforts (1 vaccine per day per 1, 000 population) could be sufficient to save 125, 839 lives in Canada during the current year 2021. We would like to point out that our study assumes that the vaccine is perfectly safe without any short or long term side-effects. This study has been conducted independently at arm’s length from vaccine manufacturers, using the available data from Canada health services. This study can be easily adapted to other places in the world.

## 1 Introduction

The dramatic spread of the novel severe acute respiratory syndrome coronavirus 2 (SARS-CoV-2) [18, 8] and the terrifying rise of the number of infected people since the first case reported in December 2019 in Wuhan [28], prompt the World Health Organization to declare the outbreak a Public Health Emergency of international concern in January 2020 [30] and a pandemic on March 11, 2020 [29]. Like worldwide countries [36, 37], Canada was not spared from the pandemic and it recorded the first presumptive case on January 25, 2020 in Toronto, Ontario. Since then, new suspected cases were increasingly reported in provinces all across the country. As of January 12, 2021, the total number of COVID-19 cases worldwide reached 91, 866, 362 with 1, 966, 276 deaths, while in Canada, the total number of COVID-19 cases reached 673, 375 with 17, 186 deaths. Before November 9, 2020, there were no approved vaccines that can provide immunity against SARS-CoV-2 infections, and only few debated pharmaceutical interventions and procedures for the treatment of COVID-19 patients were available [10, 11, 35, 16] despite no conclusive evidence of their benefit [32, 13, 15]. Non-pharmaceutical interventions (NPIs) were the only accessible strategy levers to curb the outbreak by controlling the viral transmission [9, 27]. Although there was variation among Canadian provinces or territories in the timing and stringency of NPIs implementation, these measures were the primary tools to mitigate early spread of the coronavirus disease pandemic in Canada [24, 26].

Since the publication of the genetic sequence of SARS-CoV-2, the coronavirus that causes COVID-19, in January 2020, tremendous research efforts have been devoted to the development of a vaccine against the disease. The scale of the humanitarian and economic impact of the COVID-19 pandemic has driven vaccine developers to rely on novel paradigms to accelerate the vaccine development, and the first COVID-19 vaccine candidate entered human clinical testing with unprecedented rapidity on March 16, 2020 [34]. As of April 8, 2020, the global COVID-19 vaccine R&D landscape included 115 vaccine candidates [34]. On November 9, 2020, Pfizer and BioNTech announced that their vaccine candidate against COVID-19 achieved success in the first interim analysis from Phase 3 study. They claimed that their vaccine is 90% effective in preventing COVID-19 in participants without evidence of prior SARS-CoV-2 infection in the first interim efficacy analysis. On November 16, 2021, Moderna announced their initial Phase 3 results showing a 94.5% efficacy for their COVID-19 vaccine, joining Pfizer-BioNTech as a front-runner in the global race to contain the devastating COVID-19 pandemic [25]. Canada has so far approved the Pfizer-BioNTech vaccine on December 9, 2020 and the Moderna vaccine on December 23, 2020. Both vaccines require two doses a number of weeks apart for full efficacy. First Canadians received Pfizer-BioNTech’s COVID-19 vaccine on December 14, 2020, kicking off the largest immunization campaign in the country’s history. Obviously, the vaccination campaign comes at a cost, but it is crystal clear that saving human lives is priceless. In an analysis conducted by Quadrant Health Economics for Moderna [21] it was stated that a “cost-conscious payer should be willing to pay up to $300 for a COVID-19 vaccination course with an efficacy of only 60%”. According to a study, which considers direct medical costs and productivity losses due to COVID-19, the PHICOR group at CUNY, claims that if the vaccine were to only reduce the risk of severe disease, it would be cost saving up to $200 *−* $800 for the two-dose regimen, depending on the vaccine efficacy [19]. Some politicians have also tossed the idea of paying people to get vaccinated. For instance, as reported in [39], a former US congressman suggested paying $1, 500 to every adult with a proof of vaccination, which will cost approximately $383 Billion if every adult took advantage of this program. He also claims that this program is worth the cost because it would save lives and accelerate the reopening of the economy. As per [39], the policy of paying people for COVID-19 vaccination should be adopted only in the worst case scenario where the number of voluntary vaccinated people is insufficient to promote herd immunity within a reasonable period of time [22].

A legitimate question that one may ask is what is the best vaccination strategy that healthcare policymakers should implement and how this vaccination strategy would impact the evolution of the COVID-19 pandemic. To answer this question, we developed a new mathematical prediction model referred to as the SIRV model, with Susceptible (S), Infected (I), Removed (R) and effectively-Vaccinated (V) compartments. This model, incorporating the daily-vaccination rate and the vaccine efficacy as control inputs, allows to predict the evolution of COVID-19 epidemic under the influence of the vaccination. We consider the most effective and easy-to-implement vaccination strategy, which consists of a vaccination campaign at a maximum possible daily rate over a given time-horizon. We then use our SIRV model to predict the evolution of the pandemic, for different vaccination rates and vaccine efficacy, in Canada and its most affected provinces (Ontario, Quebec, British Columbia, Alberta, Saskatchewan, and Manitoba). Note that several predictive mathematical models for epidemics, with different levels of sophistication, have been proposed or used in the literature, see for instance [1, 6, 4] and references therein. Epidemiological models incorporating the effect of vaccination have been also studied in different contexts [5, 2, 3]. Recent studies that focus on modelling and prediction of the ongoing COVID-19 pandemic can be found, for instance, in [23, 44, 17, 7, 42] but do not consider the effect of vaccination. We find it timely and important to propose a prediction approach to asses the outcomes of COVID-19 pandemic under the effect of population-wide vaccination and vaccine efficacy. Of course, our approach can be extended to other existing models to reflect the impact of the vaccination on the outcome of the predicted variables. Moreover, we believe that our proposed model is suitable for future studies on vaccine efficacy when abundant and consistent vaccination data becomes available.

## 2 Results

This work provides an assessment of the epidemiological trends of the COVID-19 pandemic in Canada and its most affected provinces, under the influence of vaccination. Our predictions are based on our newly developed mathematical model that takes into account the daily vaccination rates and vaccine efficacy as detailed in the **Methods** section. Our proposed prediction model, referred to as SIRV, is an extended version of the SIR model with a new state V representing the effectively vaccinated sub-population, and two inputs consisting of the daily-vaccination rate *u* and the vaccination efficacy *α*. The total population is partitioned into 4 sub-populations: *S*, susceptible (non-infected without immunity); *I*, infected (active cases); *R*, removed (closed cases, recovered or dead); *V*, effectively vaccinated (non-infected individuals that were effectively vaccinated, immune). In our results, the daily vaccination rates used for our epidemic predictions represent the daily vaccination rate of *fully vaccinated* individuals (*i*.*e*., non-infected individuals that received the two doses of the vaccine).

As discussed in the **Methods** section, our SIRV model has been calibrated using the data from the Public Health Agency of Canada (PHAC) from July 17, 2020 to January 8, 2021. Our projections are obtained under the assumption that there are no major changes in the NPI measures in place (*e*.*g*., face masks, social distancing, *etc*.) throughout the prediction horizon (*i*.*e*., the model parameters were assumed constant). Moreover, we would like to mention that, by the end of December 2020, provinces across Canada have implemented strict lockdowns (*e*.*g*., curfews, stay-at-home orders, travel restrictions,…*etc*) in order to help stop the spread of the coronavirus. The drop in new cases takes usually around two-to-three weeks to be reflected in the data. Therefore, our predictions (which are based on a model calibrated with data up to January 8, 2020) might be altered depending on the ongoing and the upcoming governmental decisions and lockdown measures.

We use our model to forecast the outcomes of the pandemic (up to January 2022), in Canada and its most affected provinces, under four boundary scenarios. First, we consider the worst case scenario (**Scenario 1**) where we let the pandemic take its course without vaccination. This can also be viewed as a reference scenario to the less probable situation where the vaccine is totally ineffective. It could be an alternative scenario also in the situation where there is a delay or a shortage in vaccine production and delivery [41]. In the second scenario, we assume that the vaccine is administered at a *low daily rate* corresponding to 1*/*2 daily vaccine per 1, 000 population (**Scenario 2**). This is actually (almost) the current trend in the vaccination campaigns carried out throughout Canada [https://COVID19tracker.ca/vaccinationtracker.html]. In the third scenario, we assume that the vaccine is administered at a *moderate daily rate* corresponding to one daily vaccine per 1, 000 population (**Scenario 3**). Finally, we consider the *high daily vaccination rate* scenario corresponding to two vaccines per 1, 000 population (**Scenario 4**). For each vaccination scenario, two derivative scenarios were included to assist understanding the influence of the vaccine efficacy. The first derivative scenario considers the pessimistic case where the vaccine is only 60% efficient. This is not a scenario to underestimate espe-cially with the recent studies asserting that some vaccines are less efficient than initially claimed. For instance, it is reported in [40] that Sinovac’s vaccine is barely above 50% efficacy. The second derivative scenario, however, lines up with what has been announced by major vaccine manufacturers who claim that the COVID-19 vaccine has an efficacy of 95% [38].

### 2.1 Nation-wide assessment

The parameters used in our model are given in Table 3. The results of our model calibration are depicted in Figure 9. The prediction results are summarized in Table 1 and Table 2, and illustrated in Figure 2 for the four boundary scenarios. In the latter figure, we illustrated the evolution of the active cases, closed cases, daily new cases and daily deaths, for the four boundary vaccination scenarios and the two vaccine efficacy derivative scenarios, over a time horizon that spans from January 2021 to January 2022.

**Table 1:**
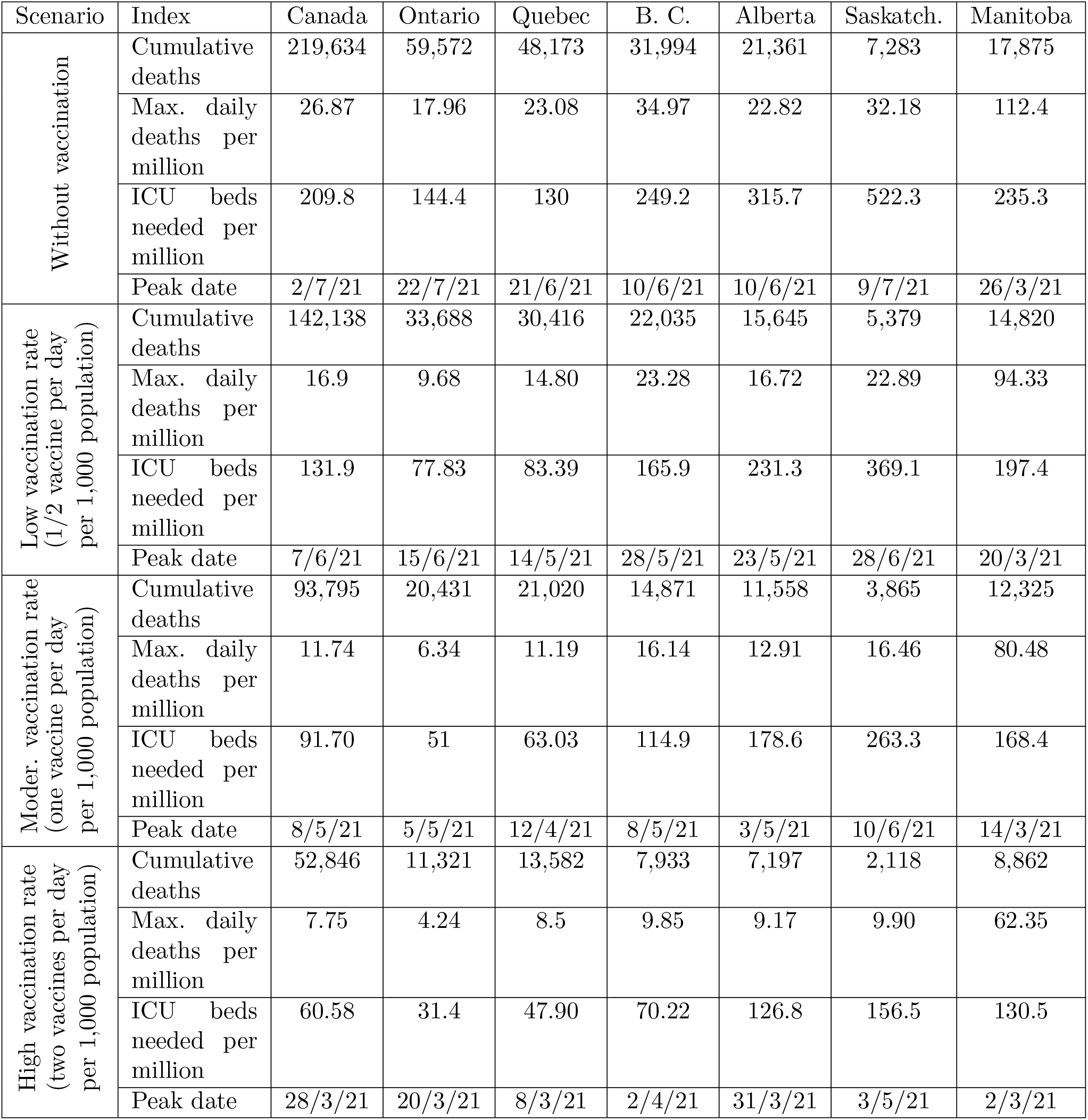
Results summary. Cumulative deaths through December 31, 2021, maximum estimated daily deaths per million population, Intensive Care Unit (ICU) beds needed per million population at the peak, and date of the maximum daily deaths (peak). These predictions are based on an optimistic vaccine efficacy of 95%.

| Scenario | Index | Canada | Ontario | Quebec | B. C. | Alberta | Saskatch. | Manitoba |
| --- | --- | --- | --- | --- | --- | --- | --- | --- |
| Without vaccination | Cumulative deaths | 219,634 | 59,572 | 48,173 | 31,994 | 21,361 | 7,283 | 17,875 |
|  | Max. daily deaths per million | 26.87 | 17.96 | 23.08 | 34.97 | 22.82 | 32.18 | 112.4 |
|  | ICU beds needed per million | 209.8 | 144.4 | 130 | 249.2 | 315.7 | 522.3 | 235.3 |
|  | Peak date | 2/7/21 | 22/7/21 | 21/6/21 | 10/6/21 | 10/6/21 | 9/7/21 | 26/3/21 |
| Low vaccination rate<br>(1/2 vaccine per day<br>per 1,000 population) | Cumulative deaths | 142,138 | 33,688 | 30,416 | 22,035 | 15,645 | 5,379 | 14,820 |
|  | Max. daily deaths per million | 16.9 | 9.68 | 14.80 | 23.28 | 16.72 | 22.89 | 94.33 |
|  | ICU beds needed per million | 131.9 | 77.83 | 83.39 | 165.9 | 231.3 | 369.1 | 197.4 |
|  | Peak date | 7/6/21 | 15/6/21 | 14/5/21 | 28/5/21 | 23/5/21 | 28/6/21 | 20/3/21 |
| Moder. vaccination rate<br>(one vaccine per day<br>per 1,000 population) | Cumulative deaths | 93,795 | 20,431 | 21,020 | 14,871 | 11,558 | 3,865 | 12,325 |
|  | Max. daily deaths per million | 11.74 | 6.34 | 11.19 | 16.14 | 12.91 | 16.46 | 80.48 |
|  | ICU beds needed per million | 91.70 | 51 | 63.03 | 114.9 | 178.6 | 263.3 | 168.4 |
|  | Peak date | 8/5/21 | 5/5/21 | 12/4/21 | 8/5/21 | 3/5/21 | 10/6/21 | 14/3/21 |
| High vaccination rate<br>(two vaccines per day<br>per 1,000 population) | Cumulative deaths | 52,846 | 11,321 | 13,582 | 7,933 | 7,197 | 2,118 | 8,862 |
|  | Max. daily deaths per million | 7.75 | 4.24 | 8.5 | 9.85 | 9.17 | 9.90 | 62.35 |
|  | ICU beds needed per million | 60.58 | 31.4 | 47.90 | 70.22 | 126.8 | 156.5 | 130.5 |
|  | Peak date | 28/3/21 | 20/3/21 | 8/3/21 | 2/4/21 | 31/3/21 | 3/5/21 | 2/3/21 |

**Table 2:** Results summary. Cumulative deaths through December 31, 2021, maximum estimated daily deaths per million population, Intensive Care Unit (ICU) beds needed per million population at the peak, and date of the maximum daily deaths (peak). These predictions are based on a pessimistic vaccine efficacy of 60%.

| Scenario | Index | Canada | Ontario | Quebec | B. C. | Alberta | Saskatch. | Manitoba |
| --- | --- | --- | --- | --- | --- | --- | --- | --- |
| Without vaccination | Cumulative deaths | 219,634 | 59,572 | 48,173 | 31,994 | 21,361 | 7,283 | 17,875 |
|  | Max. daily deaths per million | 26.87 | 17.96 | 23.08 | 34.97 | 22.82 | 32.18 | 112.4 |
|  | ICU beds needed per million | 209.8 | 144.4 | 130 | 249.2 | 315.7 | 522.3 | 235.3 |
|  | Peak date | 2/7/21 | 22/7/21 | 21/6/21 | 10/6/21 | 10/6/21 | 9/7/21 | 26/3/21 |
| Low vaccination rate<br>(1/2 vaccine per day<br>per 1,000 population) | Cumulative deaths | 167,406 | 41,637 | 35,837 | 25,435 | 17,570 | 6,044 | 15,882 |
|  | Max. daily deaths per million | 19.9 | 11.96 | 17.1 | 27.03 | 18.66 | 25.98 | 100.5 |
|  | ICU beds needed per million | 155.3 | 77.83 | 96.33 | 192.5 | 258.2 | 421.6 | 210.3 |
|  | Peak date | 18/6/21 | 1/7/21 | 28/5/21 | 3/6/21 | 30/5/21 | 1/7/21 | 22/3/21 |
| Moder. vaccination rate<br>(one vaccine per day<br>per 1,000 population) | Cumulative deaths | 126,653 | 29,140 | 27,277 | 19,848 | 14,410 | 4,937 | 14,109 |
|  | Max. daily deaths per million | 15.17 | 8.48 | 13.55 | 21.01 | 15.53 | 20.93 | 90.31 |
|  | ICU beds needed per million | 118.4 | 51 | 76.34 | 149.6 | 214.8 | 339.6 | 189.04 |
|  | Peak date | 30/5/21 | 3/6/21 | 5/5/21 | 23/5/21 | 18/5/21 | 22/6/21 | 19/3/21 |
| High vaccination rate<br>(two vaccines per day<br>per 1,000 population) | Cumulative deaths | 77,940 | 16,696 | 18,139 | 12,278 | 10,010 | 3,254 | 11,232 |
|  | Max. daily deaths per million | 10.18 | 5.48 | 10.14 | 13.75 | 11.55 | 14.07 | 74.64 |
|  | ICU beds needed per million | 79.48 | 44 | 57.12 | 97.94 | 159.9 | 228.4 | 156.2 |
|  | Peak date | 25/4/21 | 19/4/21 | 31/3/21 | 27/4/21 | 23/4/21 | 29/5/21 | 11/3/21 |

**Table 3:** Model parameter estimates for the most affected provinces in Canada. The model fitting is carried out using the official reported data covering the period between July 17, 2020 and January 8, 2021.

| Parameter | Canada | Ontario | Quebec | B. C. | Alberta | Saskatchewan | Manitoba |
| --- | --- | --- | --- | --- | --- | --- | --- |
| Basic re-production number $\mathfrak{R}_0$ | 1.25 | 1.20 | 1.19 | 1.27 | 1.32 | 1.44 | 1.26 |
| Infection rate $\beta(\times 10^{-2})$ | 9.76 | 10.67 | 10.81 | 11.25 | 8.36 | 7.10 | 18.03 |
| Removal rate $\gamma(\times 10^{-2})$ | 7.81 | 8.89 | 9.06 | 8.83 | 6.34 | 4.93 | 14.26 |
| Mortality rate $\mu(\times 10^{-2})$ | 1.64 | 1.4 | 1.96 | 1.59 | 1.14 | 1.13 | 3.35 |
| Population number $N(\times 10^6)$ | 38 | 14.7 | 8.6 | 5.15 | 4.4 | 1.18 | 1.38 |
| RMSE $(\times 10^{-4})$ | 4.50 | 3.87 | 5.90 | 5.86 | 15 | 13 | 32 |

**Figure 1:**
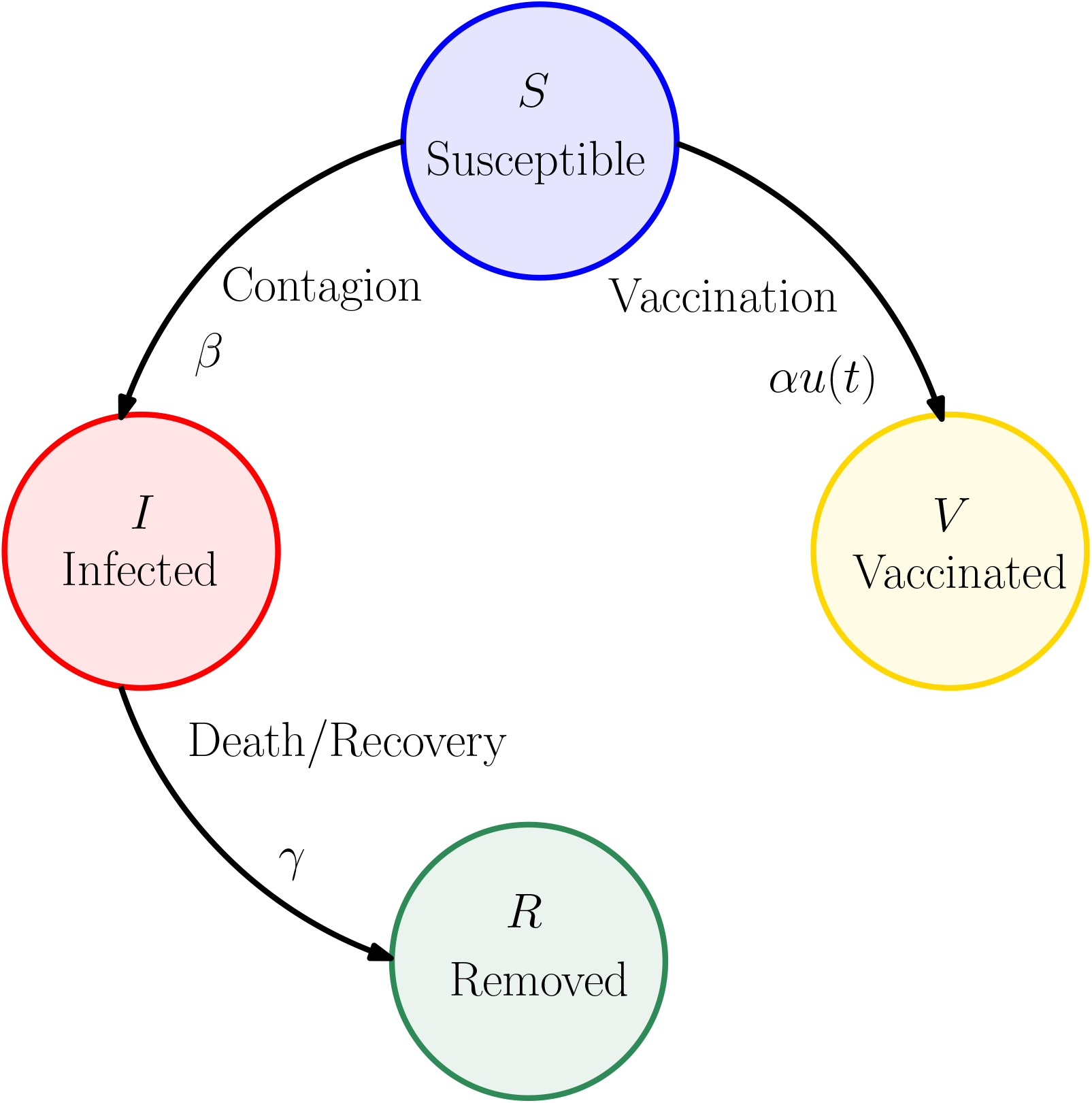
The SIRV model. A schematic graph representing the interactions between the different stages of infection in the proposed mathematical SIRV model with vaccination: *S*, susceptible (uninfected without immunity); *I*, infected (active cases); *R*, removed (closed cases, recovered or dead); *V*, effectively vaccinated (non-infected that have been effectively vaccinated, immune). The parameter *β* represents the rate of infection, *γ* is the rate of removal, and *α* represents the vaccine efficacy. The control variable *u*(*t*) represents the daily vaccination rate.

**Figure 2:**
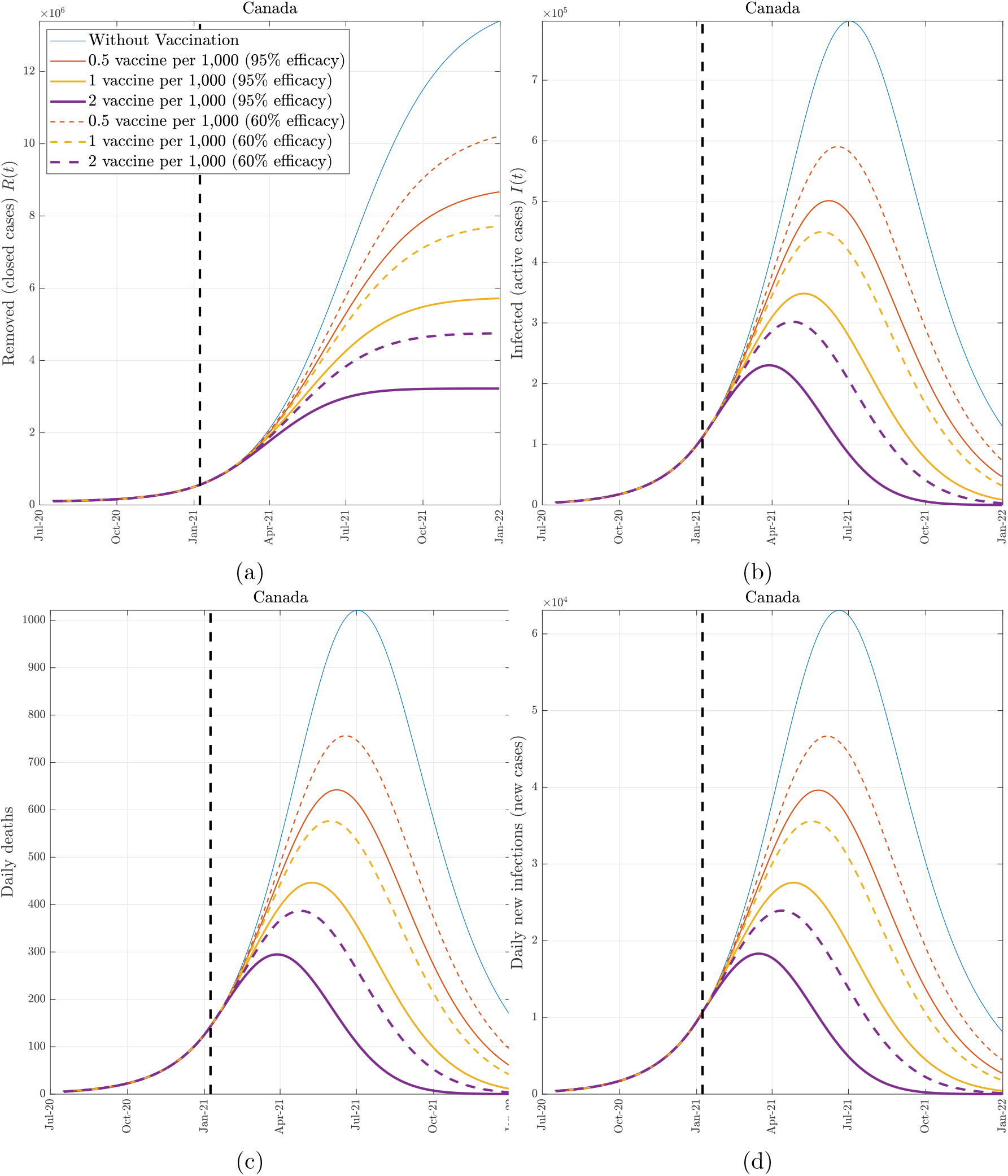
The effect of vaccination (Canada). COVID-19 epidemic evolution in Canada predicted by the proposed model from January 8, 2021 to December 31, 2021. The colored (solid and dashed) lines represent the four boundary scenarios. We consider two derivative scenarios for the vaccine efficacy: an optimistic scenario at 95% efficacy (solid lines) and a pessimistic scenario at 60% efficacy (dashed lines). The dashed vertical line identifies January 8, 2021.

**Figure 3:**
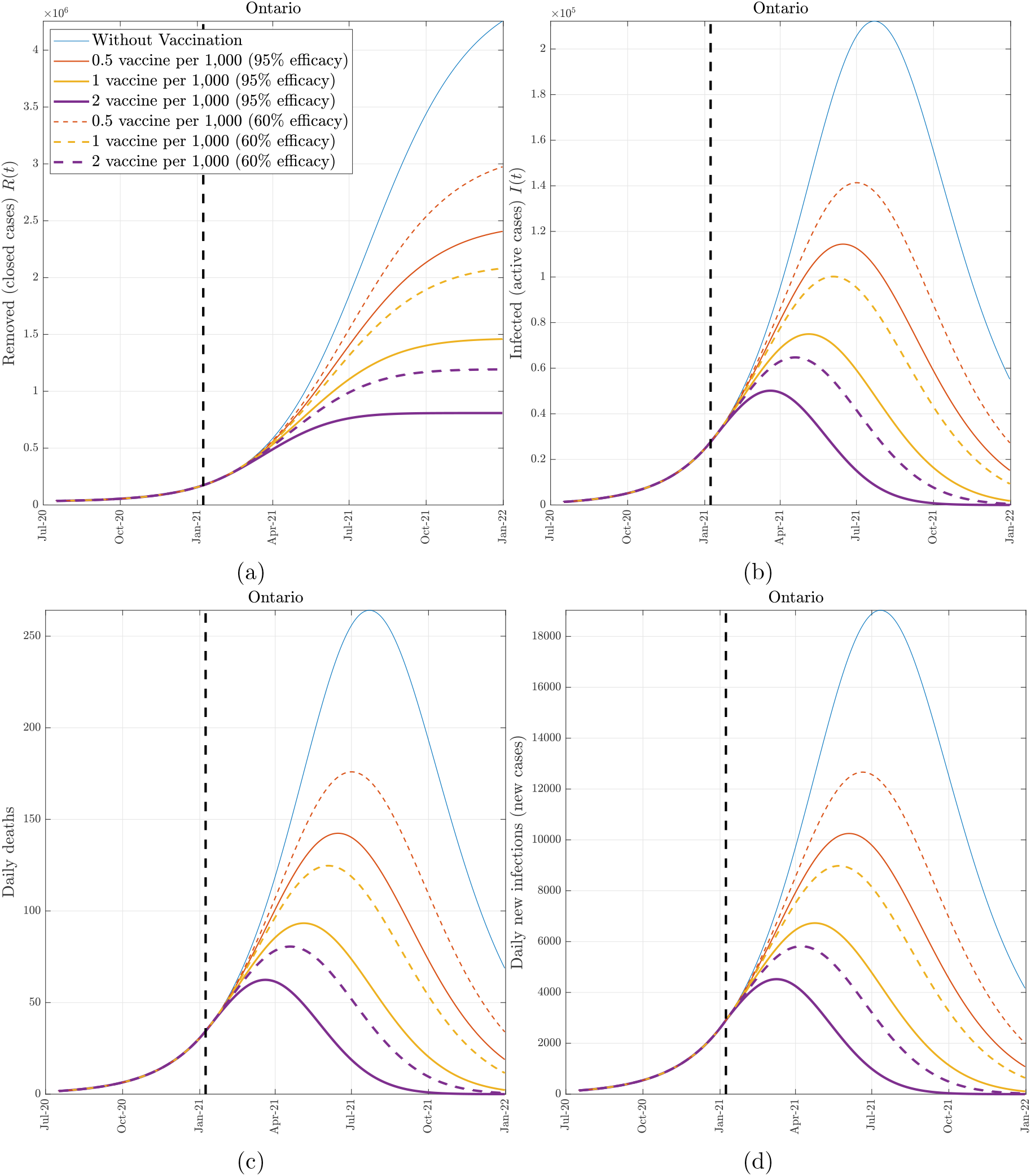
The effect of vaccination (Ontario). COVID-19 epidemic evolution in Ontario predicted by the proposed model from January 8, 2021 to December 31, 2021. The colored (solid and dashed) lines represent the four boundary scenarios. We consider two derivative scenarios for the vaccine efficacy: an optimistic scenario at 95% efficacy (solid lines) and a pessimistic scenario at 60% efficacy (dashed lines). The dashed vertical line identifies January 8, 2021.

**Figure 4:**
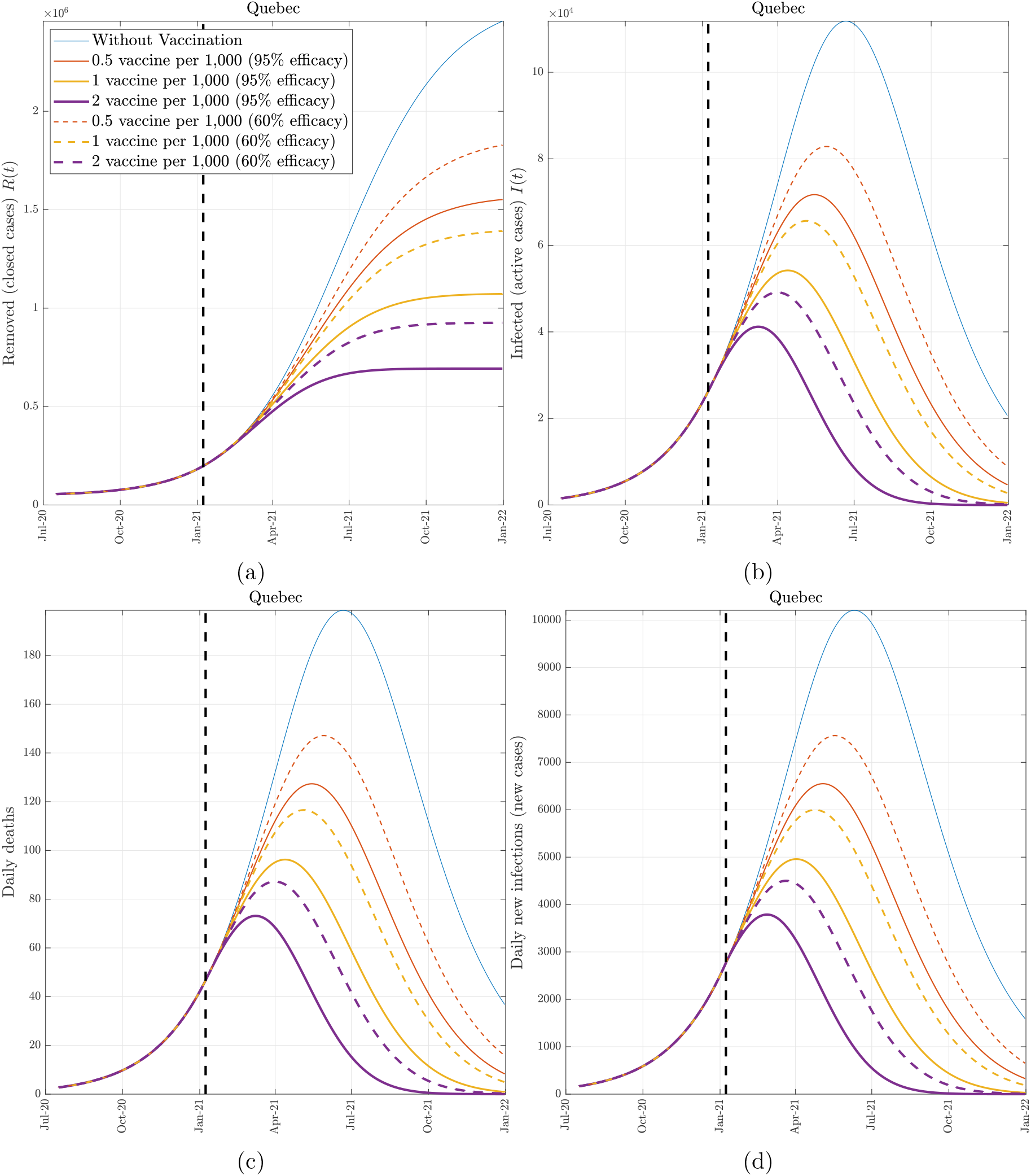
The effect of vaccination (Quebec). COVID-19 epidemic evolution in Quebec predicted by the proposed model from January 8, 2021 to December 31, 2021. The colored (solid and dashed) lines represent the four boundary scenarios. We consider two derivative scenarios for the vaccine efficacy: an optimistic scenario at 95% efficacy (solid lines) and a pessimistic scenario at 60% efficacy (dashed lines). The dashed vertical line identifies January 8, 2021.

**Figure 5:**
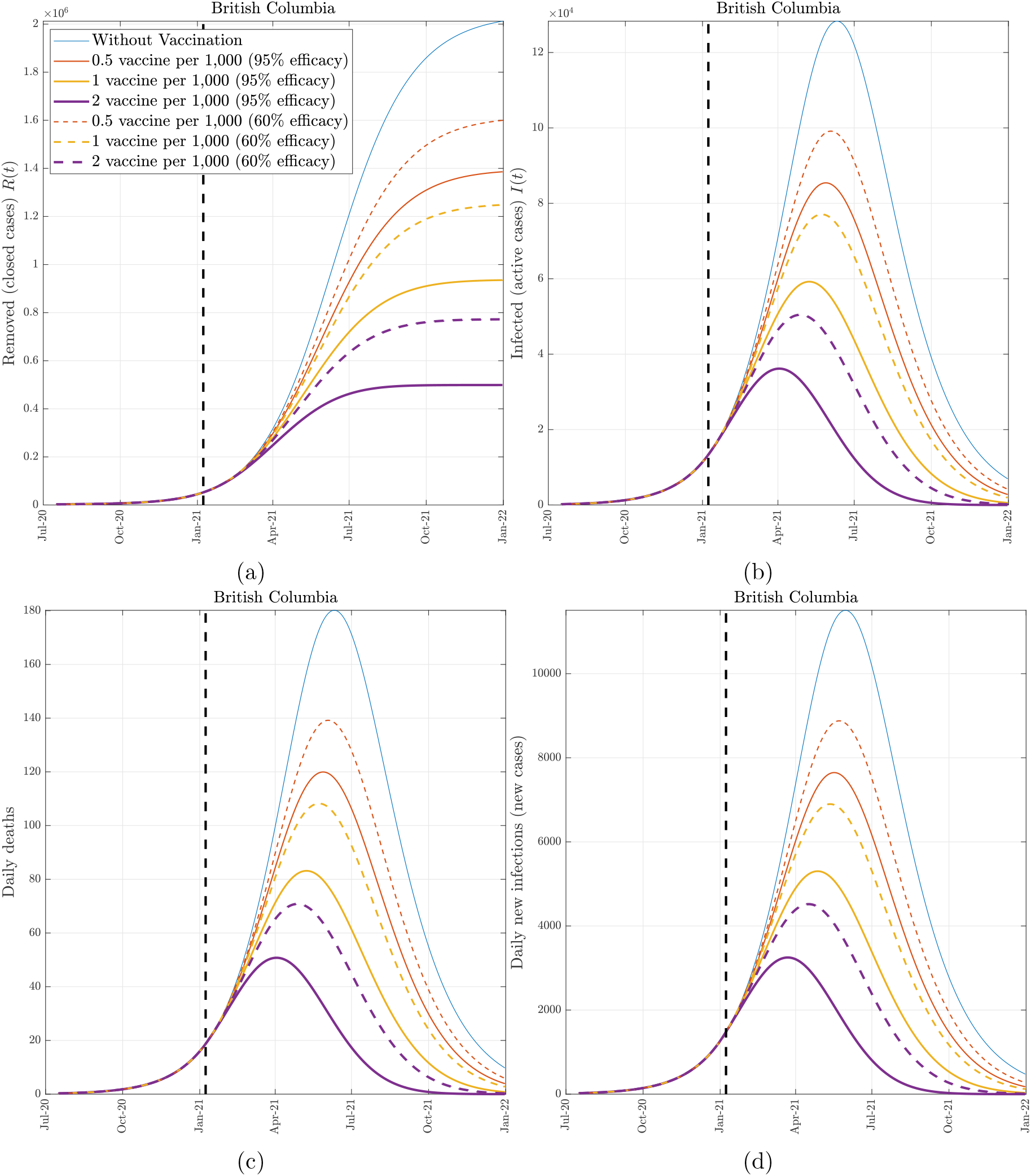
The effect of vaccination (British Columbia). COVID-19 epidemic evolution in B.C. predicted by the proposed model from January 8, 2021 to December 31, 2021. The colored (solid and dashed) lines represent the four boundary scenarios. We consider two derivative scenarios for the vaccine efficacy: an optimistic scenario at 95% efficacy (solid lines) and a pessimistic scenario at 60% efficacy (dashed lines). The dashed vertical line identifies January 8, 2021.

**Figure 6:**
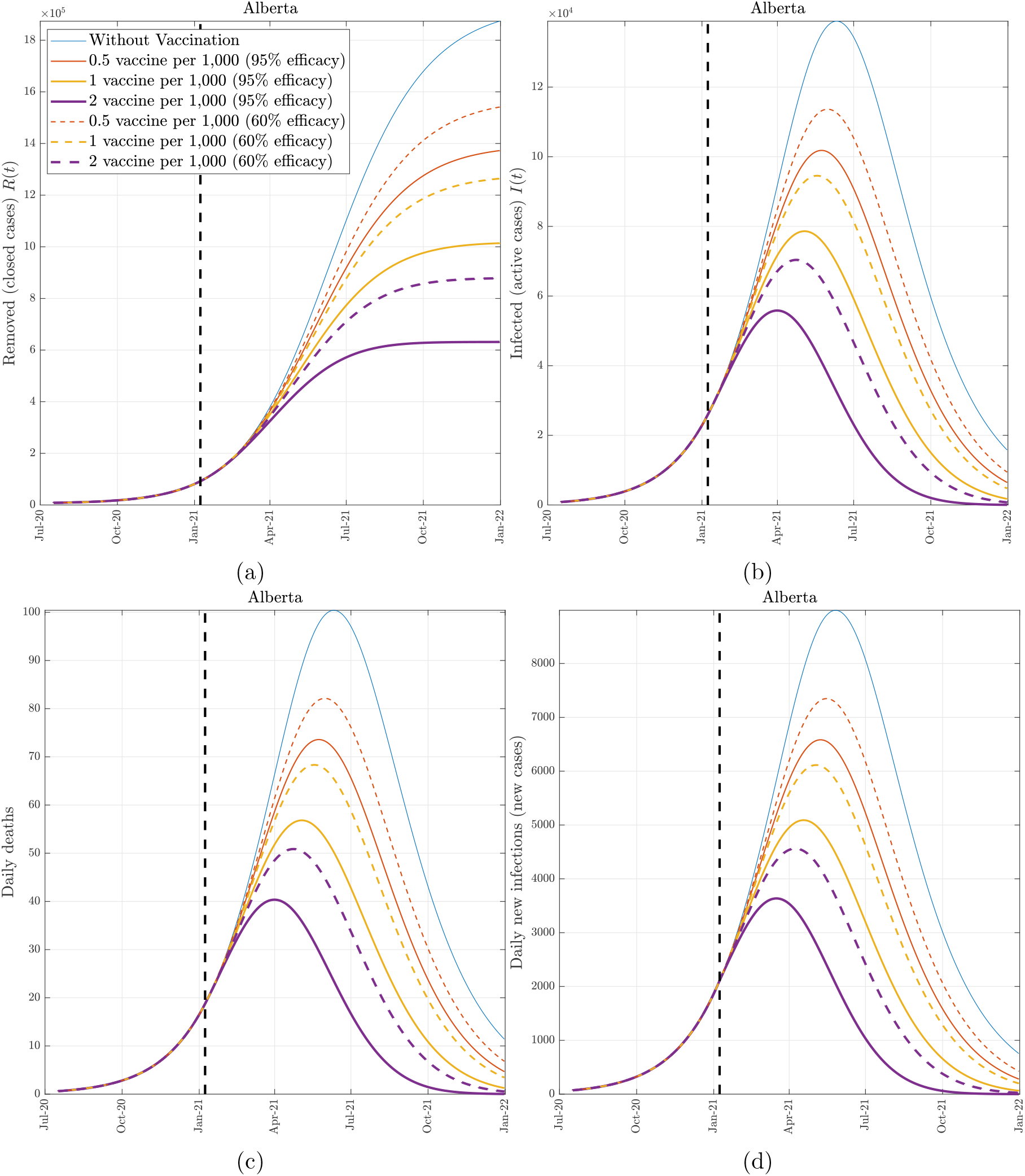
The effect of vaccination (Alberta). COVID-19 epidemic evolution in Alberta predicted by the proposed model from January 8, 2021 to December 31, 2021. The colored (solid and dashed) lines represent the four boundary scenarios. We consider two derivative scenarios for the vaccine efficacy: an optimistic scenario at 95% efficacy (solid lines) and a pessimistic scenario at 60% efficacy (dashed lines). The dashed vertical line identifies January 8, 2021.

**Figure 7:**
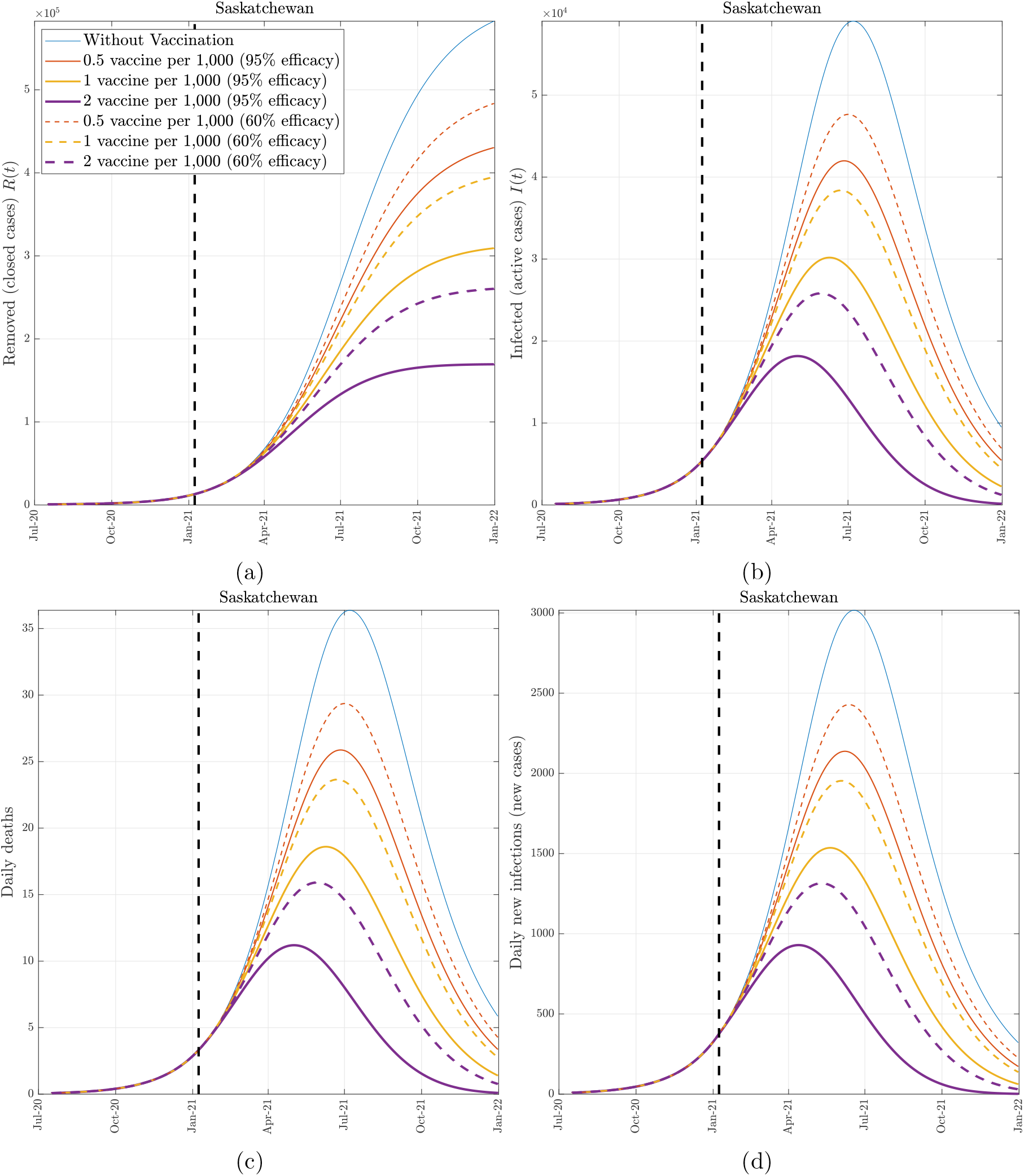
The effect of vaccination (Saskatchewan). COVID-19 epidemic evolution in Sakatchewan predicted by the proposed model from January 8, 2021 to December 31, 2021. The colored (solid and dashed) lines represent the four boundary scenarios. We consider two derivative scenarios for the vaccine efficacy: an optimistic scenario at 95% efficacy (solid lines) and a pessimistic scenario at 60% efficacy (dashed lines). The dashed vertical line identifies January 8, 2021.

**Figure 8:**
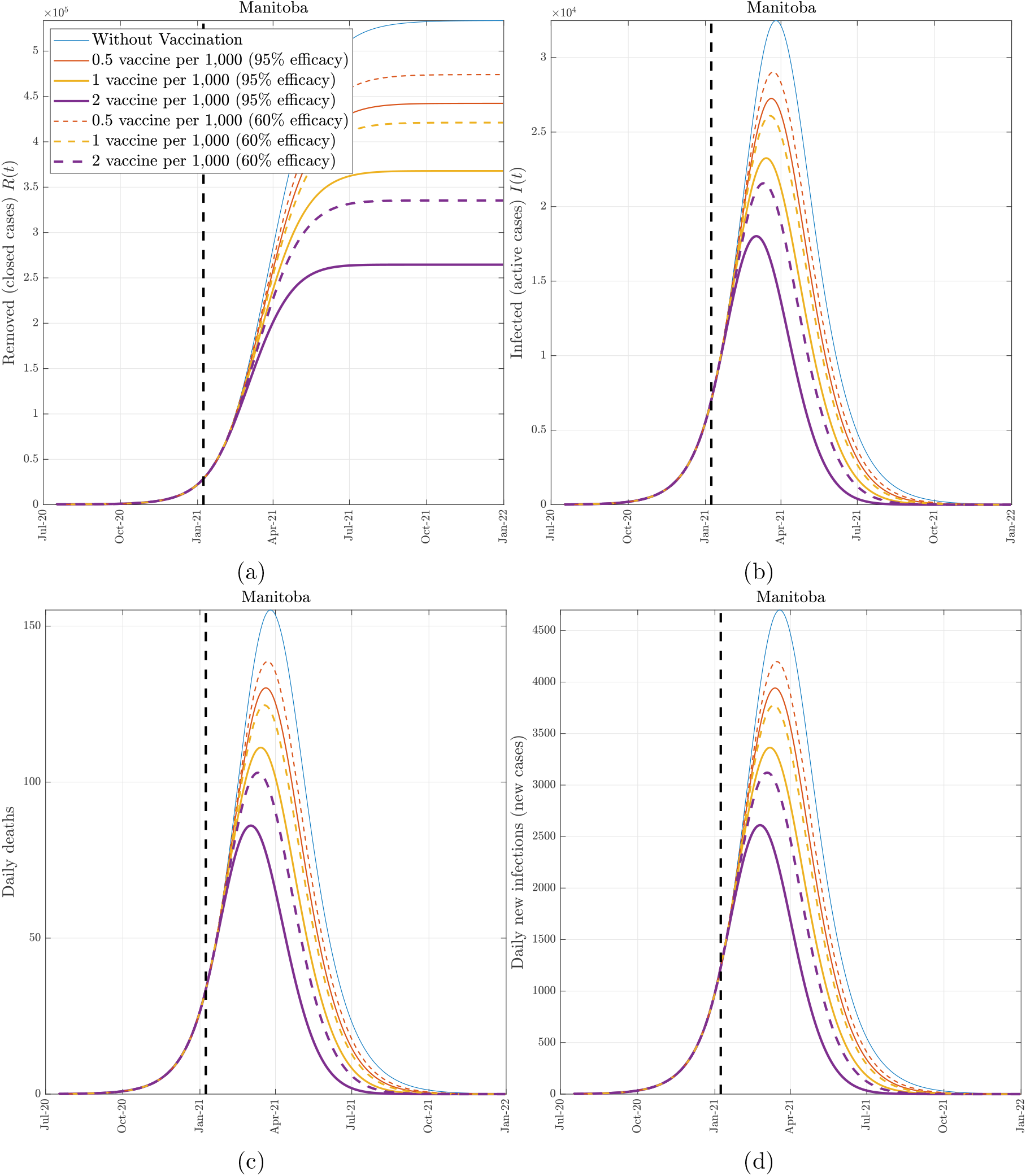
The effect of vaccination (Manitoba). COVID-19 epidemic evolution in Manitoba predicted by the proposed model from January 8, 2021 to December 31, 2021. The colored (solid and dashed) lines represent the four boundary scenarios. We consider two derivative scenarios for the vaccine efficacy: an optimistic scenario at 95% efficacy (solid lines) and a pessimistic scenario at 60% efficacy (dashed lines). The dashed vertical line identifies January 8, 2021.

**Figure 9:**
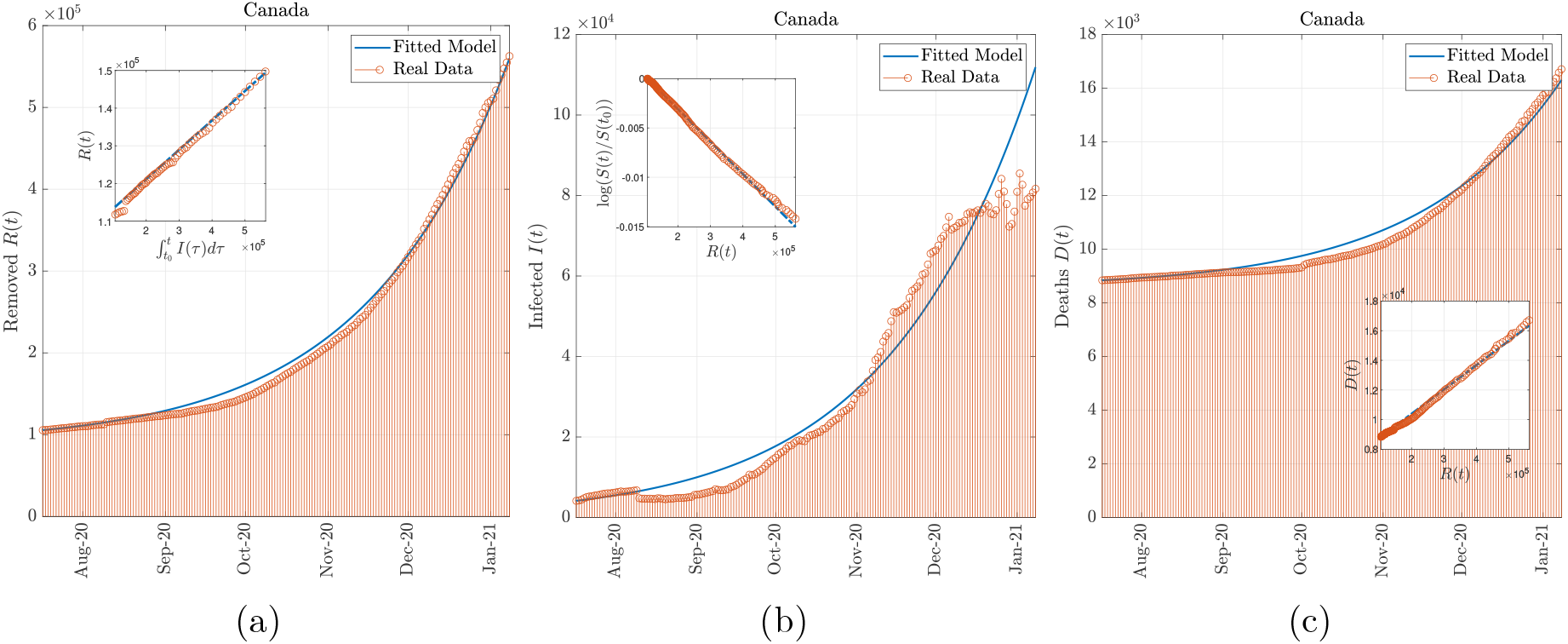
Model simulation compared to real data (Canada). Comparison between the official data (red dots histogram) and the results with the calibrated SIR model (blue line). Panel (a): number of reported removed cases (deaths and recovered). Panel (b): number of reported infected. Panel (c): number of cumulative deaths. The real data covers the period between July 17, 2020 and January 8, 2021.

#### Scenario 1 (no vaccination)

As shown in the **Methods** section, the current nation-wide basic reproduction number is ℜ_0_ = 1.25. At this pace, without any vaccination, our model predicts that the COVID-19 pandemic in Canada would last until January 2023 and by January 2022 more than 219, 000 lives will be lost. The pandemic peak is expected to occur in early July 2021 where the number of active cases would hit 797, 240, the number of daily infections would reach 63, 063, the number of daily deaths would exceed a 1, 000, and the number of needed ICU beds would be around 7, 972 (*∼* 210 beds per 1 million population). This threatening number of patients that need to be ICU-hospitalized at the same time might saturate the Canadian health system and would worsen even more the mortality burdens (more deaths).

#### Scenario 2 (low vaccination rate)

At the daily vaccination rate of 1*/*2 vaccine per 1, 000 population, which seems to match the current trend in Canada, and 95% (resp. 60%) vaccine efficacy, our model predicts that 142, 138 (resp. 167, 406) lives will be lost by the end of the year; thus saving around 77, 496 (resp. 52, 228) lives compared to Scenario 1. For an optimistic vaccine efficacy of 95%, the pandemic peak would occur in early June 2021, where the number of active cases would hit 50, 1220, the number of daily infected individuals would reach 39, 626, the number of daily deaths would approach 642, and the number of needed ICU beds would be around 5, 222. In the pessimistic scenario of 60% efficacy, the peak would occur two weeks later with 46, 668 daily new infections, 756 daily deaths, and 5, 901 ICU beds needed.

#### Scenario 3 (moderate vaccination rate)

At the daily vaccination rate of 1 vaccine per 1, 000 population, which corresponds to the double of the current vaccination trend in Canada, and 95% (resp. 60%) vaccine efficacy, our model predicts that 125, 839 (resp. 92, 981) lives could be saved by the end of the year as a result of such a consistent vaccination campaign. Moreover, with this moderate vaccination rate and 95% vaccine efficacy, the peak in new infections and new deaths would be recorded by early May 2021 where the number of active cases would hit 348, 460, the number of daily infected individuals would reach 35, 574, the number of daily deaths would approach 446, and the number of needed ICU beds would be around 3, 485. If the vaccine is only 60% efficient, we will witness the peak three weeks later with a daily death toll of 576 victims and nearly 4, 500 seriously infected patients in the ICU.

#### Scenario 4 (high vaccination rate)

At the daily vaccination rate of 2 vaccines per 1, 000 population, which corresponds to four times the current trend in Canada, our model predicts that more than 52, 846 lives will be lost by the end of the year if the vaccine is 95% efficient whereas we will have around 77, 940 cumulative deaths if the vaccine is only 60%. In both (pessimistic and optimistic) scenarios, the amount of lives that would be saved compared to the baseline scenario without vaccination is substantial. Moreover, with 95% efficacy, the pandemic peak would occur three months earlier (*i*.*e*., in late March 2021) where the number of new cases would reach 18, 297, the number of daily deaths would approach 295, and the number of needed ICU beds would be around 2, 302. A lower 60% vaccine efficacy would add up another 91 victims to the aforementioned daily death toll at the peak (which would occur in late April 2021) and an additional 718 ICU beds will be required to nurse the critically infected COVID-19 patients.

We would like to recall that the infection rate is tightly dependent on the safety measures implemented such as lock-downs, social distancing, face covering, hygiene, quarantine,…,*etc*. Our predictions are based on the assumption that the current infection rate is constant and, therefore, the aforementioned results could be much worse if these measures are to be weakened even in the presence of the vaccine.

### 2.2 Assessment across provinces

Our model has been calibrated for the most affected provinces in Canada (*i*.*e*., Ontario, Quebec, British Columbia, Alberta, Saskatchewan, Manitoba), and the obtained parameters are given in Table 3. The data fitting plots are given Figures 10–15. It should be pointed out that, although taken from the same source, the data collected from the provinces of Ontario, Quebec and British Columbia where more coherent that the data collected from the provinces of Alberta, Saskatchewan and Manitoba. This is reflected in our model fitting accuracy represented by the RMSE values shown in Table 3, where we can clearly see that the worst model fitting is the one obtained for the province of Manitoba.

**Figure 10:**
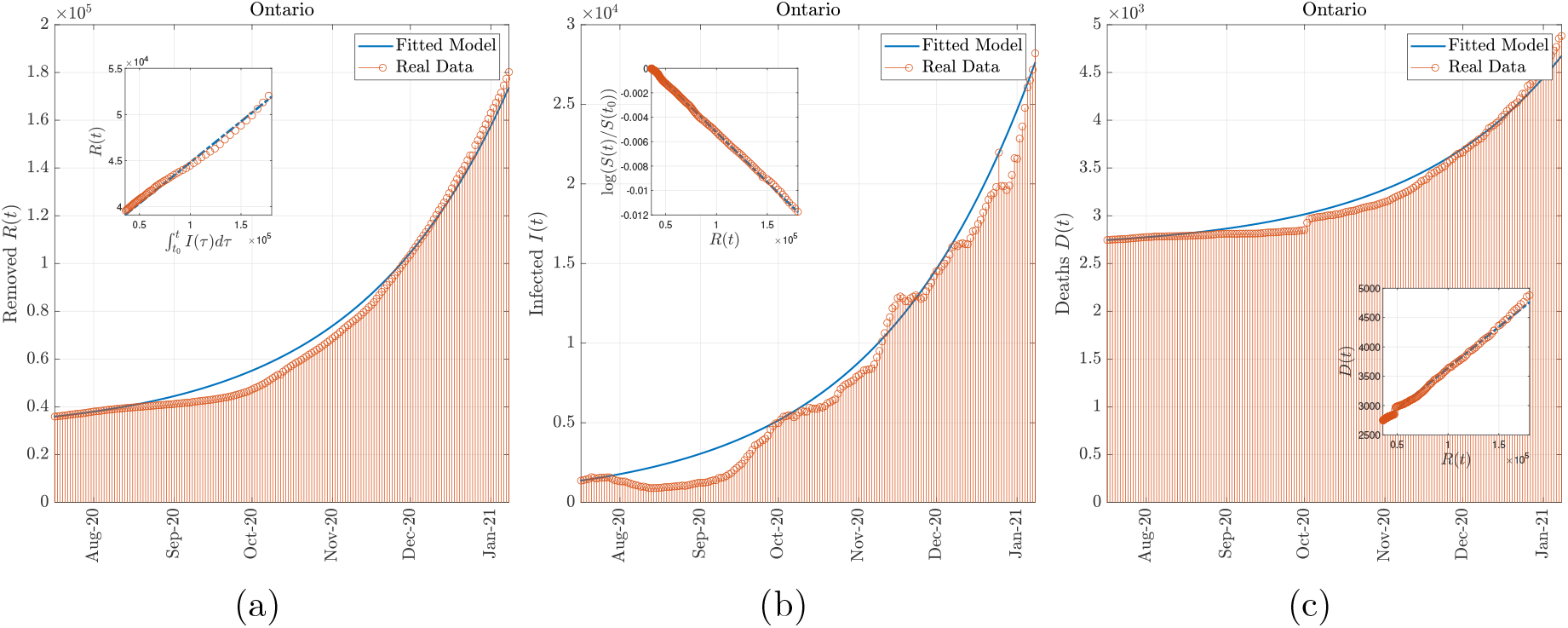
Model simulation compared to real data (Ontario). Comparison between the official data (red dots histogram) and the results with the calibrated SIR model (blue line). Panel (a): number of reported removed cases (deaths and recovered). Panel (b): number of reported infected. Panel (c): number of cumulative deaths. The real data covers the period between July 17, 2020 and January 8, 2021.

**Figure 11:**
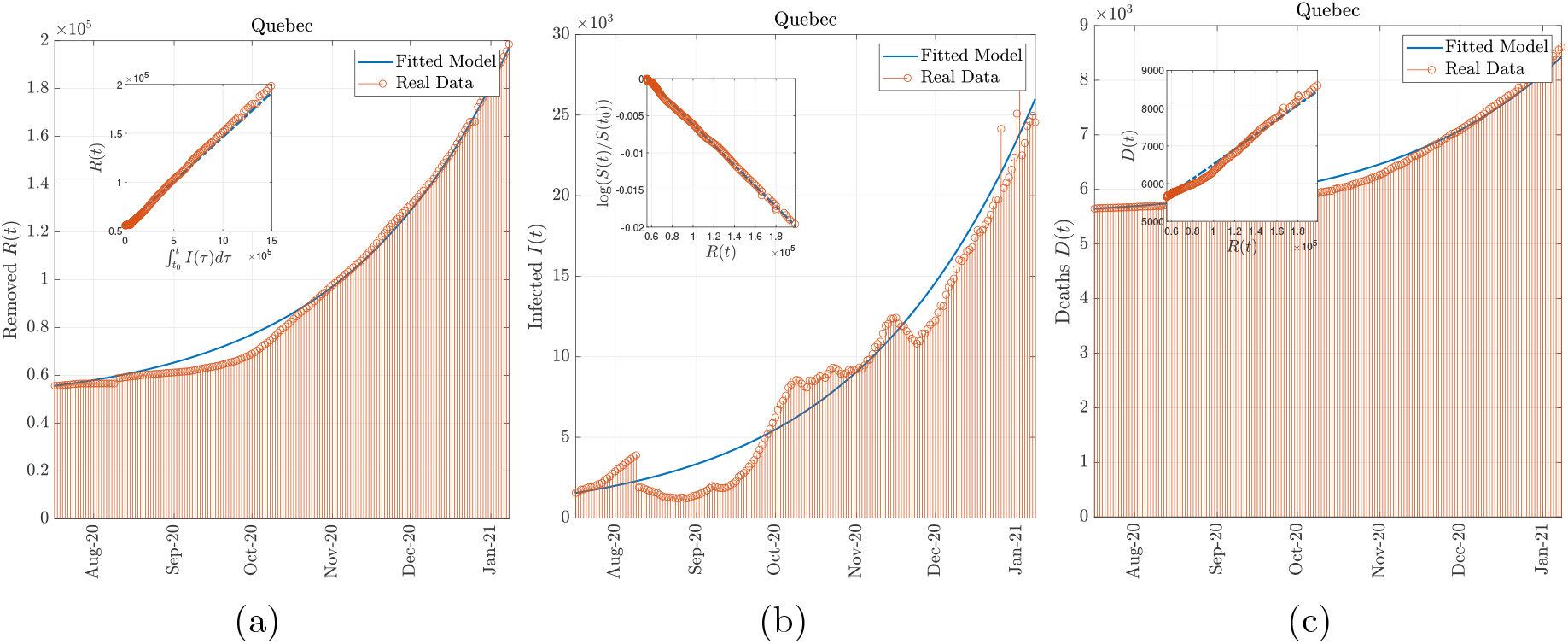
Model simulation compared to real data (Quebec). Comparison between the official data (red dots histogram) and the results with the calibrated SIR model (blue line). Panel (a): number of reported removed cases (deaths and recovered). Panel (b): number of reported infected. Panel (c): number of cumulative deaths. The real data covers the period between July 17, 2020 and January 8, 2021.

**Figure 12:**
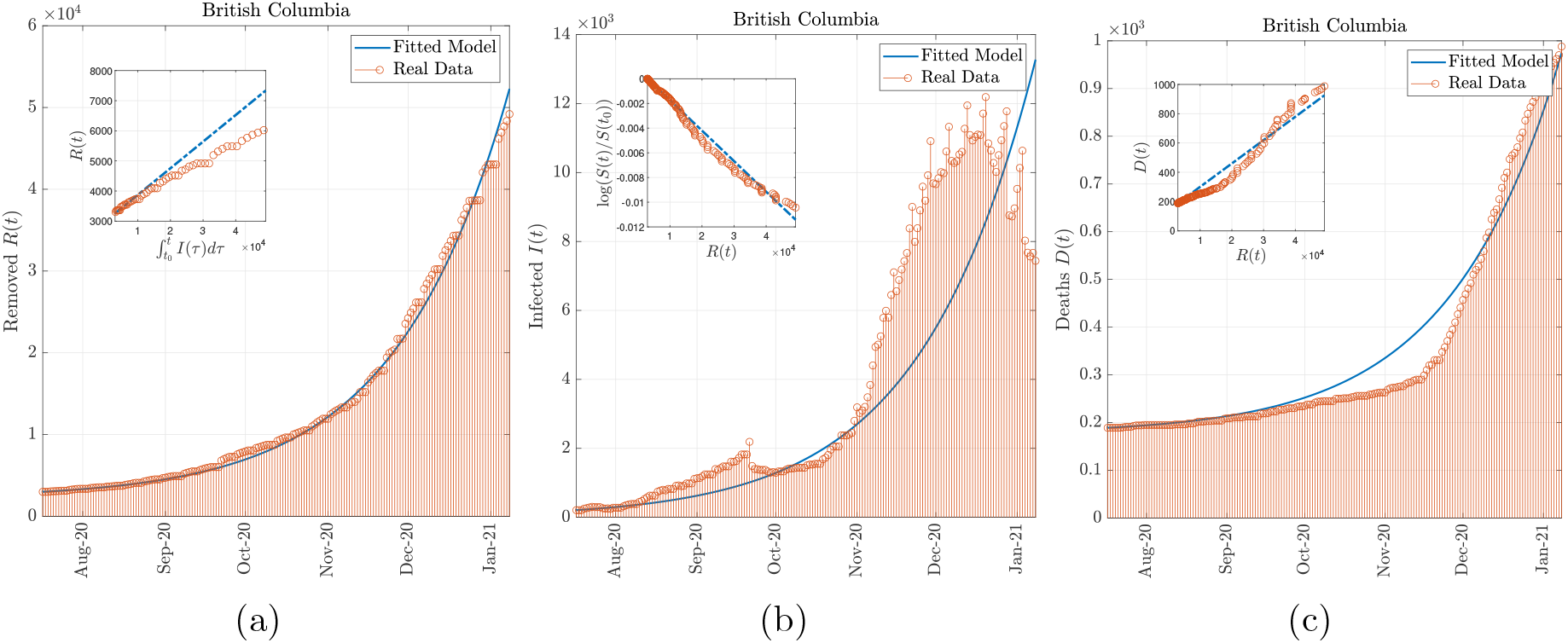
Model simulation compared to real data (British Columbia). Comparison between the official data (red dots histogram) and the results with the calibrated SIR model (blue line). Panel (a): number of reported removed cases (deaths and recovered). Panel (b): number of reported infected. Panel (c): number of cumulative deaths. The real data covers the period between July 17, 2020 and January 8, 2021.

**Figure 13:**
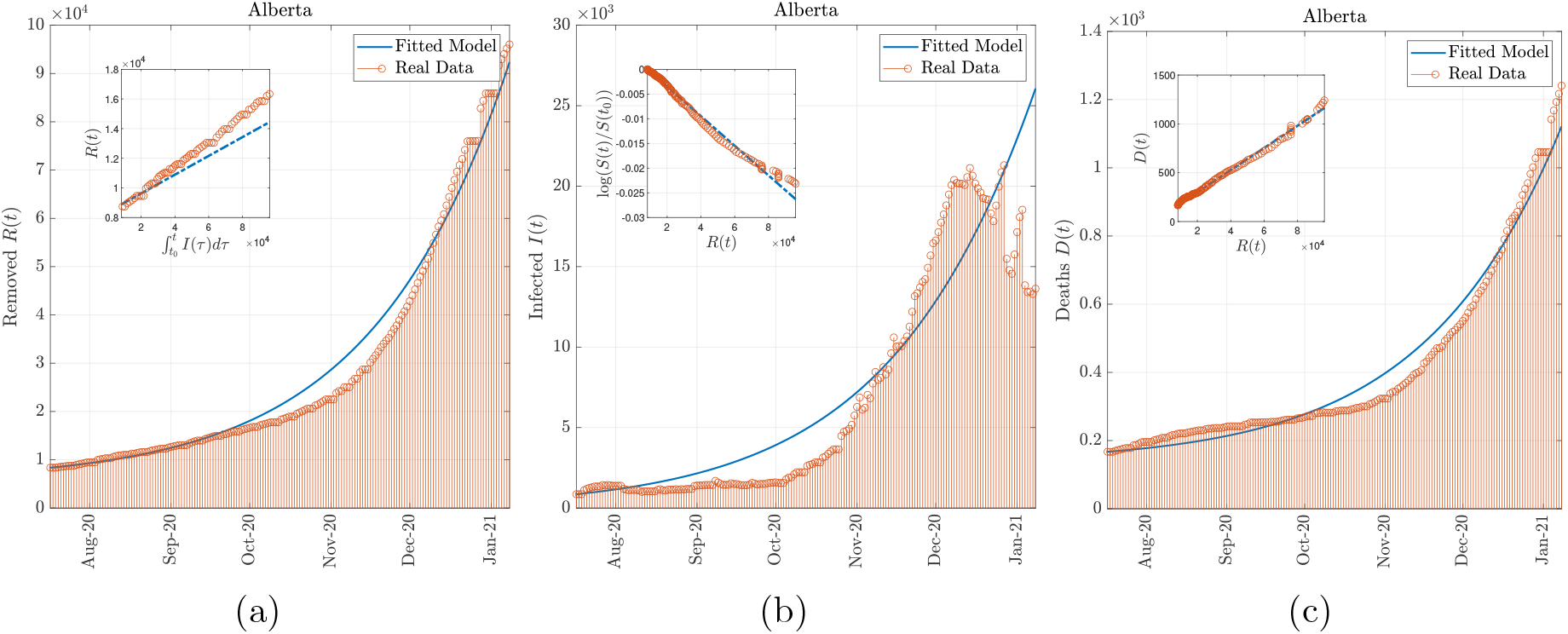
Model simulation compared to real data (Alberta). Comparison between the official data (red dots histogram) and the results with the calibrated SIR model (blue line). Panel (a): number of reported removed cases (deaths and recovered). Panel (b): number of reported infected. Panel (c): number of cumulative deaths. The real data covers the period between July 17, 2020 and January 8, 2021.

**Figure 14:**
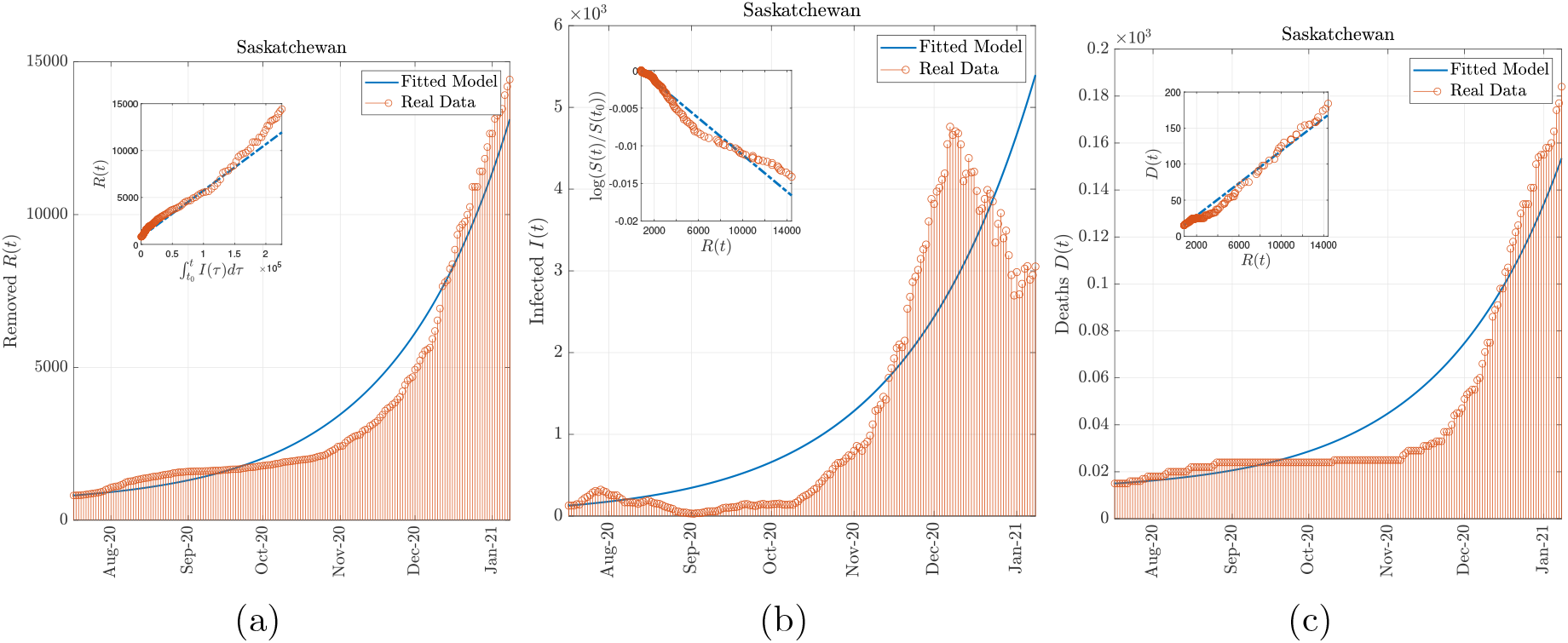
Model simulation compared to real data (Saskatchewan). Comparison between the official data (red dots histogram) and the results with the calibrated SIR model (blue line). Panel (a): number of reported removed cases (deaths and recovered). Panel (b): number of reported infected. Panel (c): number of cumulative deaths. The real data covers the period between July 17, 2020 and January 8, 2021.

**Figure 15:**
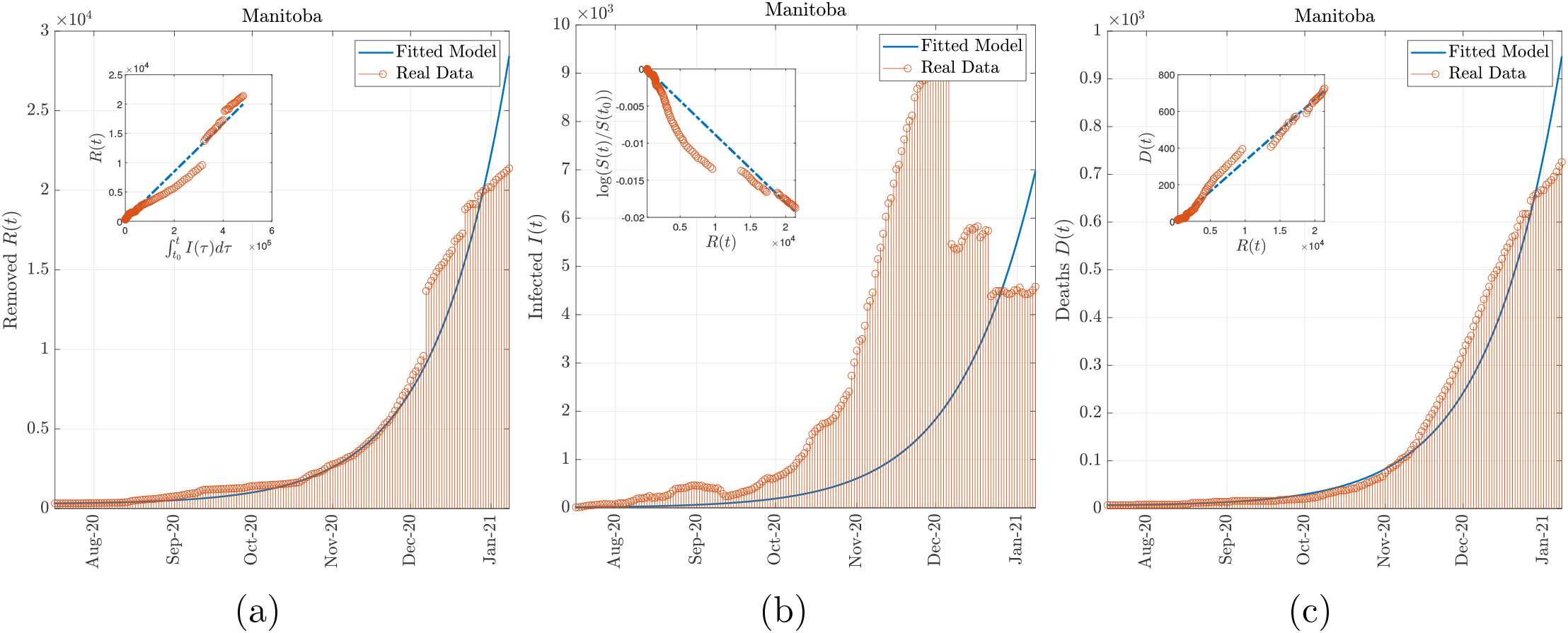
Model simulation compared to real data (Manitoba). Comparison between the official data (red dots histogram) and the results with the calibrated SIR model (blue line). Panel (a): number of reported removed cases (deaths and recovered). Panel (b): number of reported infected. Panel (c): number of cumulative deaths. The real data covers the period between July 17, 2020 and January 8, 2021.

Based on our model, we provide for each of the above mentioned provinces, some prediction results up to January 2022. We focus on the evolution of the active cases, closed cases, daily new cases and daily deaths, under different vaccination scenarios assuming a 95% or 60% vaccine efficacy. The most important pandemic indicators are extracted from our simulations and summarized in Table 1. According to the obtained results, the highest basic reproduction number and the lowest removal rate are recorded in Saskatchewan (ℜ_0_ = 1.44, *γ* = 4.93%). The highest mortality rate is recorded in Manitoba (*µ* = 3.35%) and the lowest mortality rates are recorded in Saskatchewan (*µ* = 1.13%) and Alberta (*µ* = 1.14%). The results provided for the province of Manitoba have to be taken with precaution since data corresponding to this province does not seem to be as coherent and reliable as the data from other provinces.

#### Scenario 1 (no vaccination)

Our model projections reveal that, without vaccination, the COVID-19 will continue to impose massive burdens of morbidity and mortality in all Canadian provinces. The heavy victim toll, caused by COVID-19 in Canada, would be unevenly distributed between the provinces across the country as shown in Table 1 and Table 2. The highest maximum daily deaths per million will be recorded in Manitoba (112.4) at the pandemic peak which will occur in late March 2021. The lowest maximum daily deaths per million will be recorded in Ontario (17.96) at the pandemic peak which will occur in late July 2021. The highest demand for ICU beds needed at the pandemic peak will be recorded in Saskatchewan (522.3 beds per million) in early to mid July 2021. This may be due to the fact that this province has the highest reproduction number and the lowest removal rate among all the provinces. The lowest ICU beds needed at the peak time will be recorded in Quebec (130 beds per million) in late June 2021. This may be due to the relatively low reproduction number and relatively high removal rate of this province.

#### Scenario 2 (low vaccination rate with 95% vaccine efficacy)

At a daily vaccination rate 1*/*2 vaccine per 1, 000 population, our models predicts better outcomes than the scenario without vaccinations. The trends remain similar in terms of the provinces that will have the highest and lowest burdens in terms of the maximum daily mortality and ICU beds needed at the peak times. In this scenario, the cumulative deaths that will be recorded, up to the end of this year, will toll 33, 688 in Ontario (43.4% saved lives with respect to Scenario 1) and 30, 416 in Quebec (36.9% saved lives with respect to Scenario 1).

#### Scenario 3 (Moderate vaccination rate with 95% vaccine efficacy)

At a daily vaccination rate of 1 vaccine per 1, 000 population, our models predicts better outcomes than scenario 2. The trends remain similar in terms of the provinces that will have the highest and lowest burdens in terms of the maximum daily mortality and ICU beds needed at the peak times. In this scenario, the cumulative deaths that will be recorded, up to the end of this year, will toll 20, 431 in Ontario (65.7% saved lives with respect to Scenario 1) and 21, 020 in Quebec (56.4% saved lives with respect to Scenario 1).

#### Scenario 4 (High vaccination rate with 95% vaccine efficacy)

At a daily vaccination rate of 2 vaccines per 1, 000 population, our models predicts better outcomes the previous scenarios. The trends remain similar in terms of the provinces that will have the highest and lowest burdens in terms of the maximum daily mortality and ICU beds needed at the peak times. In this scenario, the cumulative deaths that will be recorded, up to the end of this year, will toll 11, 321 in Ontario (81% saved lives with respect to Scenario 1) and 13, 582 in Quebec (71.8% saved lives with respect to Scenario 1).

### 2.3 Recommendations

Our prediction results, combined with the available data about the COVID-19 epidemic in Canada, suggest that mass vaccination at a maximum possible daily rate is urgent, necessary and cost-effective. Our findings confirm that population-wide vaccination is effective at rapidly ending the COVID-19 pandemic. Federal and provincial health agencies need to be well prepared for the surge in ICU beds needed, as per our findings, at the pandemic peak. The relatively high mortality rate in the province of Manitoba and the relatively high infection rate (inducing high demand in ICU beds *per capita* at the pandemic peak) in Saskatchewan, need to be seriously looked at by health authorities. Moreover, even in the presence of a vaccine that is quickly administered and highly efficient, our predictions do not recommend the weakening of the currently implemented safety measures. In fact, the model predictions provided in this study do not consider reduced availability of medical care due to the healthcare system reaching or even surpassing its capacity [20], which is likely going to happen at the peak of the epidemic if we do not act accordingly, resulting in worst outcomes than what our model predicts. Hence, the current adopted measures are vital to contain the pandemic and reduce the number of deaths and cannot be relieved. As new variants of the disease are appearing [43, 14] and the efficiency of the vaccines is being tested and evaluated, it is paramount to maintain the strict safety measures in place at least throughout this year.

## 3 Methods

### 3.1 SIRV model with vaccination effect

One of the earliest and well-studied mathematical models for the prediction of human-to-human propagation of infectious diseases across population is the SIR model [1]. This model relies on the time-evolution of three state variables: susceptible (S), infected (I) and removed (R). In this work, we develop a new SIRV model which extends the standard SIR model by adding the effectively vaccinated population *V* (*t*) as a state variable and the daily-vaccination rate *u*(*t*) and the vaccine efficacy *α ∈* (0, 1] as control inputs. This model relies on the principle that the susceptible population rate at time *t* will, instantaneously, decrease by *αu*(*t*) corresponding the efficacy-weighted vaccination rate. Of course, this model can be refined further by incorporating the delay between the time the vaccine was administered and the time where the vaccine will provide the expected protection and the announced efficacy. This addition was not deemed necessary at this point in time since its impact on our results and conclusions will be insignificant.

The extended SIR model with vaccination rate and vaccine efficacy inputs is depicted in Figure 1 and can be mathematically described as follows:

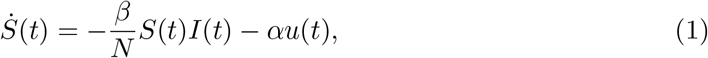

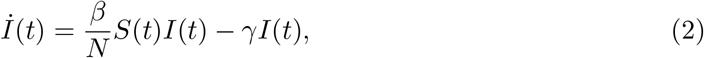

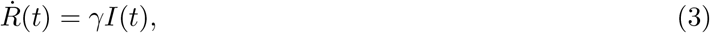

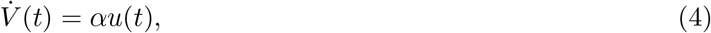

where the model state variables and parameters are defined as:

- *S*(*t*): represents the susceptible sub-population at time *t*. These are individuals not yet infected with the disease at time *t* and do not have an immunity against a potential infection.
- *I*(*t*): represents the infected sub-population at time *t* (a.k.a. active cases). These are individuals who are currently infected with the disease and are capable of spreading the disease to those in the susceptible stage.
- *R*(*t*): represents the removed sub-population at time *t* (a.k.a. closed cases). These are individuals who have been infected and then removed from the disease, either due to being recovered or dead.
- *V* (*t*): represents the effectively vaccinated sub-population at time *t*. These are individuals who have received the vaccine and have gained full immunity against the infection. Note that not all vaccinated people become necessarily immune.
- *β*: denotes the infection (transmission) rate. It represents the number of contacts *per capita* per unit time, multiplied by the probability of disease transmission in a contact between a susceptible and an infectious individual. This parameter can be modified, for example, by social-distancing and lockdowns.
- *γ*: denotes the removal rate which is the fraction of infected sub-population that is leaving the infection stage, at time *t*, to enter the removed stage. The quantity 1*/γ* represents the average infectious period.
- *α*: denotes the vaccine efficacy. It represents the ratio between the number of individuals who have received the vaccine and acquired full immunity against the disease and the total number of vaccinated sub-population.
- *u*(*t*): denotes the daily vaccination rate. It represents the number of vaccines administered at time *t*.

By November 6, 2020, there were only 17 reasonably well demonstrated cases of SARS-CoV-2 reinfection worldwide confirmed by RT-PCR and viral sequencing [33] which suggests that the reinfection rate value is negligible and can be omitted [31]. Therefore, our model assumes reasonably that the recovered cases are immune and cannot be reinfected. Moreover, our model does not take into account vital dynamics (demographic births and deaths) as these are assumed slowly varying. The total number of population remains constant, *i*.*e*.,

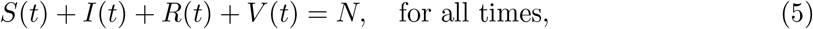

where *N* denotes the total number of the studied population. We assume that the cumulative number of deaths at a given time *t*, denoted *D*(*t*), is a fraction of the removed sub-population, *i*.*e*.,

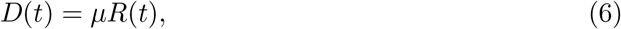

where *µ* represents the so-called mortality rate. The dynamics of the infectious class depends on the so-called *basic reproduction number*, defined by ℜ_0_ := *β/γ*, which measures the transmission potential of the disease. It represents the average number of infections caused by a single infection in a population where everyone is susceptible. Typically, a basic reproduction number that is greater than 1 would cause a proper epidemic outbreak. Our proposed model, on top of being useful in terms of studying the effect of daily-vaccination rates on the COVID-19 pandemic, shows a great potential for vaccine-efficacy determination in the future once a sufficient vaccination data is available. This can be done by fitting the SIRV model to COVID-19 data (including daily vaccination rates) to determine the vaccine efficacy *α*.

### 3.2 Fitting the model to COVID-19 epidemic data in Canada

To fit our model and infer the corresponding model parameters, we used real data provided by the Public Health Agency of Canada (PHAC) from January 30, 2020 to January 8, 2021. However, the collected data was not very reliable and consistent in the early stages of the COVID-19 pandemic due to different logistic factors as well as the fluctuations in the measures taken by the government (social distancing restrictions, quarantine, testing,…*etc*.) during the first weeks-to-months of the pandemic. Nevertheless, we have observed that the data became more consistent after July 17, 2020 as people got used to the transmission-barrier measures (face covering, disinfection, social distancing, *etc*.) and the governmental regulations. Canada has administered the first COVID-19 vaccine (by Pfizer-BioNTech) on December 14, 2020 and since then more than 260, 000 individuals have received the COVID-19 vaccine (up to January 8, 2021) which suggests a vaccination rate of around 10, 000 administered vaccine per day. The 3 weeks data, since the start of the vaccination campaign, is not enough to determine its efficacy *α* due to the low number of samples as well as the way the vaccine is administered (two shots, three to four weeks apart). This motivated us to considered the efficacy *α* as a control input in our model. The aforementioned observations and facts have motivated us to consider data samples collected between July 17, 2020 and January 8, 2021 in the estimation of the remaining model parameters *β* and *γ* by neglecting the very few vaccinated sub-population up to January 8, 2021, *i*.*e*., assuming *u*(*t*) = 0 during this period of time.

Fitting the model (1)-(3), to the COVID-19 epidemic data consists in finding the best pair of parameters (*β, γ*) that minimizes the root-mean-square error (RMSE) between the predicted state variables and their measured values. First, we note that dividing equation (1) by (3) and integrating both sides between two time instants *t*_*s*_ (data start date) and *t*_*e*_(data end date), yields the following formula:

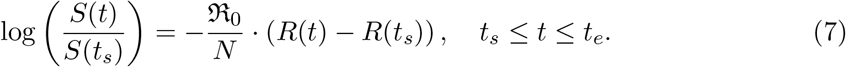

This equation shows that there is a linear relationship between log(*S*(*t*)) and the removed population *R*(*t*). This is a simple linear regression problem and is straightforward to solve using standard data fitting numerical tools. We then solve for the basic reproduction number ℜ_0_. The removal rate *γ* can be calculated using the following integral formula:

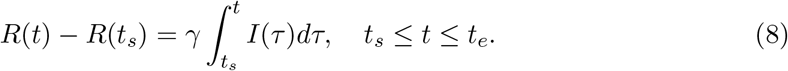

The integral in the right hand side of the above equation is calculated using the trapezoidal integration rule. The removal rate *γ* is obtained by minimizing the sum of the squares of the errors between the true values of *R*(*t*) *− R*(*t*_*s*_) (from the data) and the ones predicted by the model, *i*.*e*., the integral in (8). The infection rate *β* is obtained from *β* = ℜ_0_ *γ*.

An important challenge in tuning the model is that the initial data are affected by statistical distortion and, therefore, the model fitting process must take this problem into account. For this reason, the aforementioned obtained values of *β* and *γ* are further refined using a nonlinear programming solver that takes these values as initial conditions and searches for the best values that minimize the RMSE between the real data and the predicted samples (best-fit approach). The final estimated parameters (for Canada) are *β* = 0.0976, *γ* = 0.0781, and ℜ_0_ = 1.25 with a (normalized) RMSE = 4.5 *×* 10^*−*4^. We also perform another linear regression to determine the mortality rate *µ* from the relation in equation (6), which is found to be *µ* = 0.0164. The predicted data from the model and the real data are plotted in Fig. 9. The fitting process was also conducted for the six worst affected provinces: Ontario, Quebec, British Columbia, Alberta, Saskatchewan, and Manitoba. The parameter estimates for each of these provinces are reported in Table 3 while the comparisons between the predicted data from the model and the real data for each province are reported in Figures 10–15. The estimated population number *N* for each province is taken from Statistics Canada [12]. Finally, according to the Public Health Infobase (https://health-infobase.canada.ca/COVID-19/ epidemiological-summary-COVID-19-cases.html) 5.5% of the COVID-19 cases needed hospitilization and around 1% of them were admitted to the Intensive Care Unit (ICU). We therefore used the a rate of 1% from the active cases when calculating the ICU beds needed at the peak time, as reported in Table 1 and Table 2.

### 3.3 Vaccination strategy

Most response measures for the COVID-19 pandemic seek to “flatten the curve”, that is, to contain the growth rate of the number of cumulative infected people (active plus closed cases *R* + *I*) via a combination of testing, social distancing and lockdowns. Clearly, social distancing and lockdowns carry significant social and economic costs and they affect directly the model parameter *β* (infection rate). Our goal is to study the effect of the vaccination strategy, over a fixed period of time, on the outcome of the COVID-19 epidemic. In the case were the monetary cost is irrelevant and the main objective it to contain the COVID-19 pandemic and minimize human lives losses, it is obvious that the best strategy is to start vaccinating as soon as possible at a maximum daily rate possible. The vaccination campaign should stop only when there are no susceptible sub-population left. In other words, we propose the following daily vaccination rate

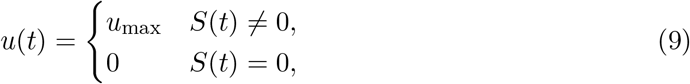

where *u*_max_ is the maximum population that can be vaccinated per day (maximum vaccination rate). Based on our proposed SIRV model (1)-(4) under the control input (9), we studied the effect of the daily vaccination rate on the evolution of the COVID-19 pandemic in Canada and its most affected provinces and the results are discussed in the **Results** section. This is done by integrating the nonlinear dynamics (1)-(4) using forward Euler method with a step size equals 1 hour (1/24 days). The initial states (on July 17, 2020) can be read from the first row of the data table in the **Source data**. The states are propagated, using the estimated parameters as discussed in Section 3.2 and shown in Fig. 9, from July 17, 2020 to January 8, 2021 under no vaccination input and the effect of vaccination is then introduced on January 8, 2021.

## Data Availability

We gathered COVID-19 epidemiological data from the Public Health Infobase which is managed by the Health Promotion and Chronic Disease Prevention Branch (HPCDPB) of the Public Health Agency of Canada (PHAC).

https://health-infobase.canada.ca/COVID-19/

## 4 Acknowledgements

This research work is supported by the National Research Council of Canada, under the grants NSERC-DG RGPIN-2020-04759 and NSERC-DG RGPIN-2020-0627.

Our thought are with those who lost their loved ones in this pandemic, and our gratitude and respects go to all the front-line workers across Canada and worldwide for keeping us safe and keeping the world running. Although our results predict that the upcoming year might be another year of challenge, we also predict that, by the end of the year, there is a real hope of seeing the end of the tunnel.

## 5 Author contributions

I. H. collected and analyzed the data and participated in formulating the model and providing the interpretation of the results. S. B. proposed the model and performed the mathematical derivations, the fitting and the predictions. A. T. provided critical feedback and helped shape the research work. All authors substantially contributed in the writing and the critical analysis of the results.

## 6 Competing interests

The authors declare no competing interests

## 7 Data availability

We gathered COVID-19 epidemiological data from the Public Health Infobase (https://health-infobase.canada.ca/COVID-19/) which is managed by the Health Promotion and Chronic Disease Prevention Branch (HPCDPB) of the Public Health Agency of Canada (PHAC). Raw source data are provided with this paper.

## 8 Code availability

The code used for the predictions reported in this work is publicly available online (http://flash.lakeheadu.ca/~tayebi/SIRV_code/). Analyses were carried out using MATLAB version R2020b, Curve Fitting Toolbox version 3.5.12, Signal Processing Toolbox version 8.5, and Statistics and Machine Learning Toolbox version 12.0.

## Notes

### Competing Interest Statement

The authors have declared no competing interest.

### Author Declarations

University of Quebec in Outaouais research ethics committee (CER/UQO)

